# Suspected Adverse Drug Reactions of the Type 2 Antidiabetic Drug Class Dipeptidyl-Peptidase IV inhibitors (DPP4i): Can polypharmacology help explain?

**DOI:** 10.1101/2022.06.30.22277085

**Authors:** Lauren Jones, Alan M. Jones

**Affiliations:** Medicines Safety Research Group (MSRG), School of Pharmacy, University of Birmingham, Edgbaston, Birmingham, B15 2TT, United Kingdom

**Keywords:** Dipeptidyl-Peptidase IV Inhibitors, Pharmacokinetics, Polypharmacology, Drug-related Side effects, adverse drug reactions

## Abstract

To interpret the relationship between the polypharmacology of dipeptidyl-peptidase IV inhibitors (DPP4i) and their suspected adverse drug reaction (ADR) profiles using a national registry.

A retrospective investigation into the suspected ADR profile of four licensed DPP4i in the United Kingdom using the National MHRA Yellow Card Scheme and OpenPrescribing databases. Experimental data from the ChEMBL database alongside physiochemical (PC) and pharmacokinetic (PK) profiles were extracted and interpreted.

DPP4i show limited polypharmacology alongside low suspected ADR rates. We found minimal statistical difference between the unique ADR profiles ascribed to the DPP4i except for total ADRs (*χ*^2^; *p* <.05). Alogliptin consistently showed the highest suspected ADR rate per 1,000,000 items prescribed. Saxagliptin showed the lowest suspected ADR rate across all organ classes but did not reach statistical difference (*χ*^2^; *p* >.05). We also confirmed the Phase III clinical trial data that showed gastrointestinal and skin reactions are the most reported ADR across the class and postulated underlying mechanisms for this based on possible drug interactions.

We have proposed underlying mechanisms behind the reported suspected ADRs and their polypharmacology. The main pharmacological mechanism behind the ADRs is attributed to interactions with DPP4 activity and/or structure homologue (DASH) proteins which augment the immune-inflammatory modulation of DPP4.

## Introduction

Type II diabetes is a metabolic disorder primarily associated with insulin sensitivity causing a functional deficit (insulin resistance) which may deteriorate into reduced excretion. Diabetes mellitus is the 9^th^ leading cause of death worldwide [1] and is often a result of cardiovascular events such as myocardial infarction or stroke. In the United Kingdom (UK), diabetes is estimated to account for in the region of 10% of the National Health Service (NHS) budget.[2]

UK clinical guidance for type II diabetes recommends lifestyle advice and modification as the first-line therapy, followed by metformin as a second-line therapy.[2] Intensification of treatment comprises of “add-on” therapies (five are licensed in the UK); Dipeptidyl peptidase-4 inhibitors (DPP4i), pioglitazone, sulfonylureas (SU), sodium-glucose co-transporter 2 inhibitors (SGLT2i) and, glucagon-like peptide-1 (GLP-1) analogues, which can only be initiated under specialist care.

DPP4 is an enzyme which degrades the incretin hormones GLP-1 and glucose-dependent insulinotropic polypeptide (GIP), which serve to control glycaemic levels post-prandially by stimulating insulin synthesis and secretion from pancreatic *β*-cells and reducing glucagon secretion from *α*-cells.[3][4][5] Additionally, GIP/GLP-1 delay gastric emptying and exert central nervous system modulation, thereby increasing satiety and reducing further food intake. DPP4i increase the circulating incretin hormones levels for longer post-prandially, leading to better glycaemic control and have shown preservation of pancreatic *β*-cell function through increased cell stimulation, proliferation, differentiation, and survival[6][7][8] - theoretically this could slow, or reverse, disease progression.

DPP4i’s have preferential effects over other drug classes depending on patient factors. For example, DPP4i’s are considered weight neutral, having a negligible risk of hypoglycaemia, [4][7] whereas SU and pioglitazone have a higher risk of hypoglycaemic episodes and are associated with weight gain.[9][10][11] Pioglitazone cannot be initiated in patients with heart failure (HF) due to potential complications.[10] SGLT2is are associated with a higher risk of diabetic ketoacidosis (DKA) and limb and foot amputations.[12] GLP-1 analogues have poor oral bioavailability and are administered via injection.

Adverse drug reactions (ADR) are the unintended side effects of drugs at clinical doses for an indicated disease. ADRs can significantly impact pharmaceutical management of up to 20% of hospitalised patients’ and 25% of outpatient’s care.[13] This is associated with a significant cost; associated NHS hospitalisations equate to £380M per year.[14]

Polypharmacology is how a single drug interacts multiple targets.[15] Side effects can be a result of interactions with both the desired target and/or other targets within the body, potentially causing harm or death.[13]

The UK’s Medicines and Healthcare products Regulatory Agency (MHRA) Yellow Card Scheme [16] involves voluntary reporting from healthcare professionals and patients. It provides post-marketing surveillance of observational ADRs of UK-licensed drugs in complex real-world settings. [17] Monitoring with this spontaneous reporting system identifies further unanticipated or previously undetected ADRs and this information influences future clinical guidance. At the time of writing, there are five UK licensed DPP4i (approval date in parentheses): sitagliptin (2007); vildagliptin (2007), saxagliptin (2009), linagliptin (2011), and alogliptin (2013) (**Figure 1**).

**Figure 1.**
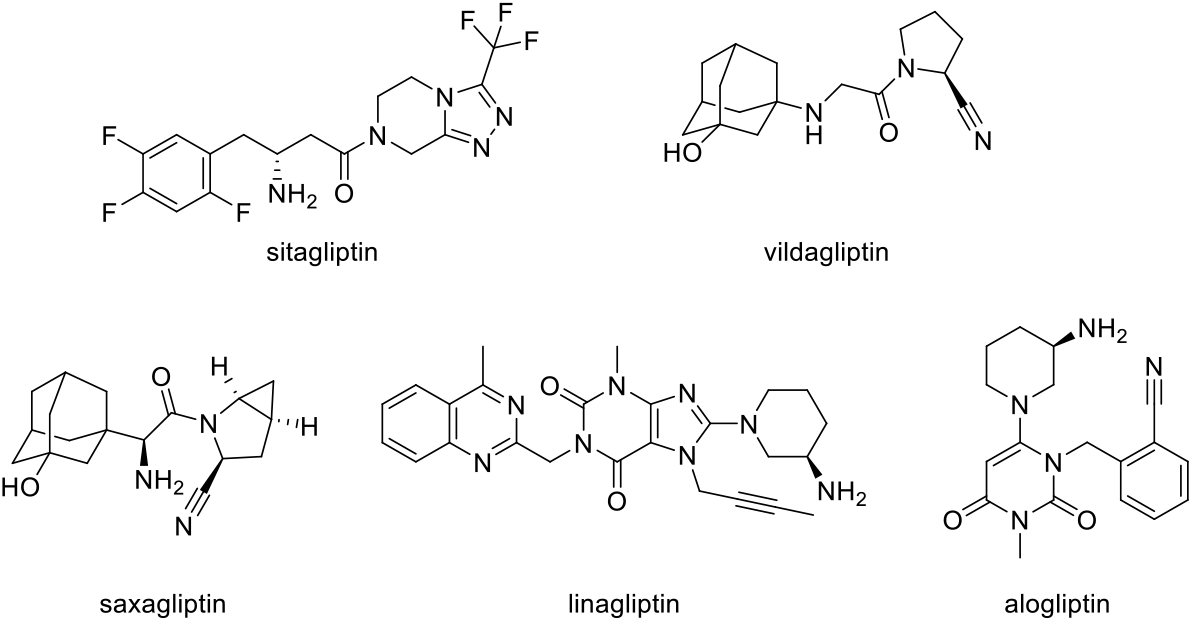
Chemical structures of the DPP4i studied.

Research linking the polypharmacology and suspected ADR profiles of DPP4i has to the best of our knowledge yet to be conducted. Investigation of the polypharmacology of each DPP4i and attributing polypharmacology to ADRs could potentially enable prospective ADR prediction in this drug class or indeed others.[18][19] Furthermore, where an ADR profile is significantly harmful, which may be attributed to its polypharmacology, this may influence clinical guidance or associated clinical decision making.

The aims of this study are to:

a. determine the unique polypharmacology of each DPP4i class member by assessing the affinity of each drug to a series of tested physiological enzymes;
b. use a national registry (Yellow Card Scheme) to determine suspected ADR signal rates relative to prescribing levels in the population;
c. run statistical analysis to ascertain which ADR signals are statistically significant and
d. attempt to establish links between the each DPP4i’s polypharmacology and unique suspected ADR profile by postulating underlying mechanisms.

The objective of this research is to ascertain the links (if any) between the polypharmacology of a DPP4i and the reported ADR profile standardised to prescribing levels.

## Methods

### Physiochemical Properties

Physiochemical (PC) properties influence the absorption, distribution, metabolism, and excretion (ADME) of a given drug and are key to determine the activity *in vivo*. These properties were compared across the DPP4i class to determine whether they influenced the pharmacology and side effect profile of each drug.

Physiochemical properties such as p*K*_*a*_ (-log_10_*K*_*a*_;where *K*_*a*_ is the acid dissociation constant), hydrogen bond donators/acceptors (HBDs/HBAs), Log_10_P (experimentally measured **P**artition coefficient of a substance between organic and aqueous environments in its neutral form) and cLog_10_P (calculated **P**artition coefficient) were data-mined from the **Ch**emical databased of bioactive molecules of the **E**uropean **M**olecular **B**iology **L**aboratory (ChEMBL), Wellcome Trust Genome Campus, Hinxton, UK[20] and molecular weight and topological polar surface area (^*t*^PSA) from PubChem.[21] Log_10_D^7.4^ (**D**istribution coefficient is the partition coefficient for an ionised compound) was calculated from the respective Log_10_P, p*K*_*a*_, and pH values. Log_10_D^7.4^ gives an indication of the distribution between aqueous and organic phase at pH = 7.4 and indicates the lipophilicity of the molecule in an ionised state. ^*t*^PSA considers the polar interactions on the surface of the molecule in deciding the conformational shape of a drug and is linked to oral absorption and blood-brain barrier (BBB) penetration.[22]

### Pharmacokinetic properties

Pharmacokinetic (PK) properties were characterised through European Medicines Agency (EMA) Summary of Product Characteristics (SPC) profiles.[23][24][25][26] PK properties included *C*_max_ (maximum plasma concentration), volume of distribution (V_d_), plasma protein binding (PPB), half-life (*t*_1/2_), renal clearance (Cl), bioavailability (%*F*), and method of clearance/excretion. Where the *C*_max_ was reported in alternative units (ng/mL), it was converted to nM to compare against various target IC_50_ values (nM) to determine if the reported value had a physiological relevance.

*p*IC_50_ was calculated as the -Log_10_(IC_50_) from the median IC_50_ value of the intended target, DPP4, and used to determine Ligand-Lipophilicity Efficiency (LLE = *p*IC_50_ – clog_10_P) which indicates the promiscuity of the molecule from the target enzyme. LLE quantifies potency with a target against lipophilicity and a lower value is associated with more toxicity (<5 is considered toxic).[27] Where parameters were not given in the resources listed, literature searches using SciFinder^®^ for the PK property + drug name were used to gather missing information[28][29][30]

### Blood Brain Barrier (BBB) penetration

The threshold of BBB penetration was assigned using; molecular weight <450 Da; neutral or basic drugs (determined by pK_a_); ^*t*^PSA <90 Å; <6 hydrogen bond donors; <2 hydrogen bond acceptors; log_10_D^7.4^ 1-3; and whether it is a P-glycoprotein (P-gp) substrate. The more BBB penetrant properties a drug possesses, the more likely a drug will cross the BBB.[31]

### Pharmacological Properties

Pharmacological target searches were completed using drug name searches on ChEMBL (accessed 29/10/2021);[20] a database that extracts and standardises bioactivity data from medicinal chemistry journals and other databases to identify biological interactions.[32][33][34] Median IC_50_ values were calculated to negate extremities of values, as IC_50_ values were tested across multiple labs at different time points. IC_50_ quantitively shows the concentration of a substance required to inhibit a protein by 50%. ChEMBL searches were refined to single protein assays (*homosapien*) to make the physiological activity as relevant as possible for human use. Those with interactions >100,000 nM were disregarded from reporting due to insignificant physiological relevance.

### Prescribing Data

The Openprescribing.net resource provided prescribing data in primary care across England from January 2017 to August 2021.[35] The prescription items (*R*_*x*_) were collated to standardise the ADRs to consider differing prescribing rates between drugs within the class. This is a different to typical pharmacovigilance studies (that don’t consider drug prescribing levels) and enables a surrogate standardisation for suspected ADRs to be more realistically compared between drug class members

### Adverse Drug Reactions (ADR) Data

ADR data was collated from the MHRA Yellow Card Scheme Interactive Drug Analysis Profiles(iDAP) [16] with dates set between January 2017 to Aug 2021 for single active constituents only. The categories reported were system organ classes; fatalities; and sub-category terms selected by a threshold ADR rate of 1.5 ADRs/1,000,000 *R*_*x*_ for at least one of the four drugs. The Yellow Card Scheme did not report ADRs for vildagliptin after Dec 2020 and therefore did not fit the scope of this research, which considered ADRs between January 2017 to August 2021. Vildagliptin was therefore excluded from further analysis due to incomplete datasets

### Statistical Analysis

Chi squared (χ^2^) analysis was performed on the standardised ADR/1,000,000 *R*_*x*_ using Microsoft Excel Version 16.55. The test was performed across all four drugs (**Table 3**) and then drug vs drug analysis (**S1**). A *p* value of <.05 was set for statistical significance.

### Ethical Considerations

All data collected was publicly available without patient identifiable information. No ethical approval or consent was required.

## Results

### Physiochemical Property Results

Chemical properties including molecular weight, ^*t*^PSA and LLE were similar across the class of DPP4i (**Table 1**). All DPP4i had LLE > 5 suggesting they are non-promiscuous inhibitors, with saxagliptin (8.82) possessing the most efficient binding and linagliptin (6.20) the least. All DPP4i, except for linagliptin, had a negative Log_10_D^7.4^, which is associated with hydrophilicity. Saxagliptin exhibited the highest potential to cross the BBB and therefore central nervous system effects may be anticipated. The remaining DPP4i studied had a low propensity to cross the BBB.

**Table 1.** Physiochemical and pharmacokinetic properties for the DPP4i studied.

|  | Alogliptin (benzoate) | Linagliptin | Saxagliptin (hydrochloride) | Sitagliptin (phosphate monohydrate) |
| --- | --- | --- | --- | --- |
| **Physiochemical Properties** |  |  |  |  |
| **MW (Da)** | 461.5 | 472.55 | 351.9 | 505.31 |
| **pK<sub>a</sub>** | 9.47 | 9.86 | 7.9 | 8.78 |
| **HB Acceptors** | 7 | 10 | 4 | 5 |
| **HB Donors** | 1 | 1 | 2 | 1 |
| **<sup>t</sup>PSA (Å<sup>2</sup>)** | 131 | 114 | 90.4 | 155 |
| **Log<sub>10</sub>D<sup>7.4</sup>** | -0.91 | 0.34 | -0.70 | -0.14 |
| **P-gp substrate** | No | Yes | No | Yes |
| **BBB penetration score** | 3 | 2 | 5 | 2 |
| **Log<sub>10</sub>P** | 0.39 | 1.15 | 1.16 | 2.02 |
| **cLog<sub>10</sub>P** | 1.16 | 2.80 | -0.08 | 1.26 |
| **IC<sub>50</sub> (nM) of DPP4 ±SD** | 5.3 ±2.60 | 1.0 ±0.66 | 3.4 ±20.4 | 18.0 ±22.6 |
| **pIC<sub>50</sub>** | 8.28 | 9.00 | 8.47 | 7.74 |
| **LLE** | 7.12 | 6.20 | 8.55 | 6.48 |
| **Pharmacokinetic Properties** |  |  |  |  |
| **Dose (mg)** | 25 OD | 5 OD | 5 OD | 100 OD |
| **F (%)** | ~100 | 30 | 75 | 87 |
| **Route** | Oral | Oral | Oral | Oral |
| **Dosage form** | Tabs | Tabs | Tabs | Tabs |
| **t<sub>1/2</sub> (h)** | 21 | 12 | 27 | 12.4 |
| **C<sub>max</sub> (nM)** | 295.8 | 11 | 68.2 | 950 |
| **Renal clearance (mL/min)** | 170 | 70 | 230 | 350 |
| **Method of excretion** | 76% in urine, 13% in faeces | 5% renal excretion, 80% in faeces | 22% faeces, 75% urine (24% as saxagliptin, 36% as metabolite) | 87% urine, 13% faeces |
| **V<sub>d</sub> (L)** | 417 | 1110 | 151 | 198 |
| **PPB (%)** | 20-30 | Concentration dependent: 75-99% | Negligible | 38 |

DPP4i given in their formulated salts. Key: BBB = blood-brain barrier; cLog<sub>10</sub>P = calculated logarithmic octanol-water Partition coefficient; HB = hydrogen bond; Da = Daltons (g/mol); Log<sub>10</sub>D<sup>7.4</sup> = logarithmic distribution coefficient at pH 7.4; MW = molecular weight; P-gp = P-glycoprotein; IC<sub>50</sub> = Half-maximal inhibitory concentration; pIC<sub>50</sub> = -log<sub>10</sub>IC<sub>50</sub>; PPB = plasma protein binding; pK<sub>a</sub> = -log<sub>10</sub>acid dissociation constant; V<sub>d</sub> = volume of distribution; <sup>t</sup>PSA = topological polar surface area; Å = Angstrom; LLE = lipophilic ligand efficiency; nM = nanomolar.

### Pharmacokinetic results

All reported drugs have once daily administration at varied doses and have similar half-lives. Notably, linagliptin hada high V_d_ which suggests a higher distribution around tissues in the body, and this may be explained by the Log_10_D^7.4^ value being the most lipophilic of the DPP4i studied. Furthermore, linagliptin had the lowest oral bioavailability, significantly higher PPB and reduced renal clearance.

### Pharmacological properties

Linagliptin had the strongest interaction with DPP4 (1.0 nM), followed by saxagliptin (3.4 nM) and alogliptin (5.3 nM). Sitagliptin showed the weakest activity with DPP4 (18 nM). Linagliptin also showed a stronger affinity for Fibroblast Activation Protein (FAP) (89 nM) as well as activity with M1 receptors (**Table 2**). Saxagliptin had inhibitory activity with FAP, but stronger affinity interactions with proteins DPP8 and DPP9. However, these interactions should not be considered in isolation; given that the *C*_max_ is considerably lower than off-target interactions, they may not be physiologically relevant. The most physiologically relevant interactions can be seen in **Table 2**.

**Table 2.**
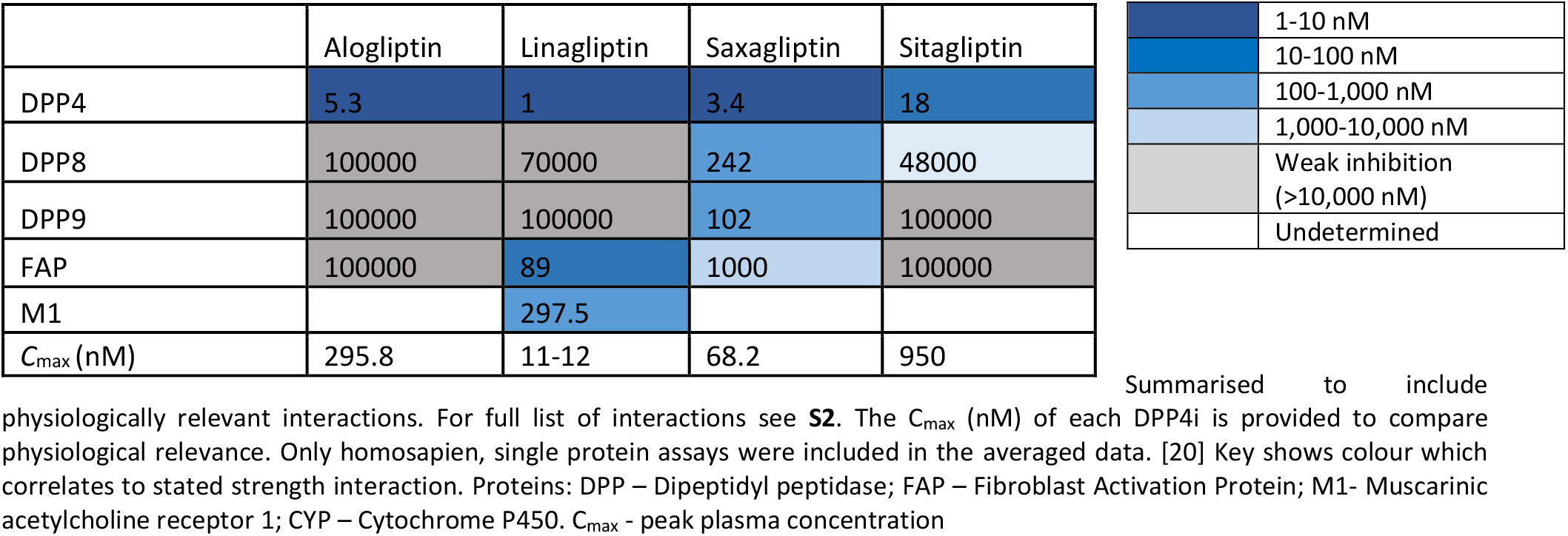
Summarised median IC_50_ values (nM) for four DPP4i against named proteins.

|  | Alogliptin | Linagliptin | Saxagliptin | Sitagliptin |
| --- | --- | --- | --- | --- |
| DPP4 | 5.3 | 1 | 3.4 | 18 |
| DPP8 | 100000 | 70000 | 242 | 48000 |
| DPP9 | 100000 | 100000 | 102 | 100000 |
| FAP | 100000 | 89 | 1000 | 100000 |
| M1 |  | 297.5 |  |  |
| C<sub>max</sub> (nM) | 295.8 | 11-12 | 68.2 | 950 |

|  |
| --- |
| 1-10 nM |
| 10-100 nM |
| 100-1,000 nM |
| 1,000-10,000 nM |
| Weak inhibition (>10,000 nM) |
| Undetermined |

Summarised to include physiologically relevant interactions. For full list of interactions see **S2**. The C<sub>max</sub> (nM) of each DPP4i is provided to compare physiological relevance. Only homosapien, single protein assays were included in the averaged data. [20] Key shows colour which correlates to stated strength interaction. Proteins: DPP – Dipeptidyl peptidase; FAP – Fibroblast Activation Protein; M1- Muscarinic acetylcholine receptor 1; CYP – Cytochrome P450. C<sub>max</sub> - peak plasma concentration

### Open Prescribing

Prescribing data shows that sitagliptin is the most prescribed DPP4i, with 11.5 million items (*R*_*x*_), followed by linagliptin (9.4 million *R*_*x*_), alogliptin (5.6 million *R*_*x*_) and saxagliptin (1.0 million *R*_*x*_). Number of prescriptions does not necessarily equate to number of patients as the drugs are available in a variety of formulated tablet strengths (**S3**) and multiple tablets may be required to reach the daily indicated dose.

### ADR results

Given the significant difference in prescribed numbers between the four drugs, the ADRs were standardised per 1,000,000 *R*_*x*_ for accurate comparison whilst mitigating the risk of misinterpreting unstandardised values.

### Total ADRs

Alogliptin had the highest total ADR rate with 98.18 per 1,000,000 Rx followed by linagliptin (69.81), sitagliptin (55.24) and saxagliptin (53.01) (**Table 3**). This result was statistically significant across the DPP4i class (**Table 3**) with a *p* <.05. Furthermore, alogliptin reached statistical difference when analysed individually against other DPP4i studied (**S1**).

**Table 3.**
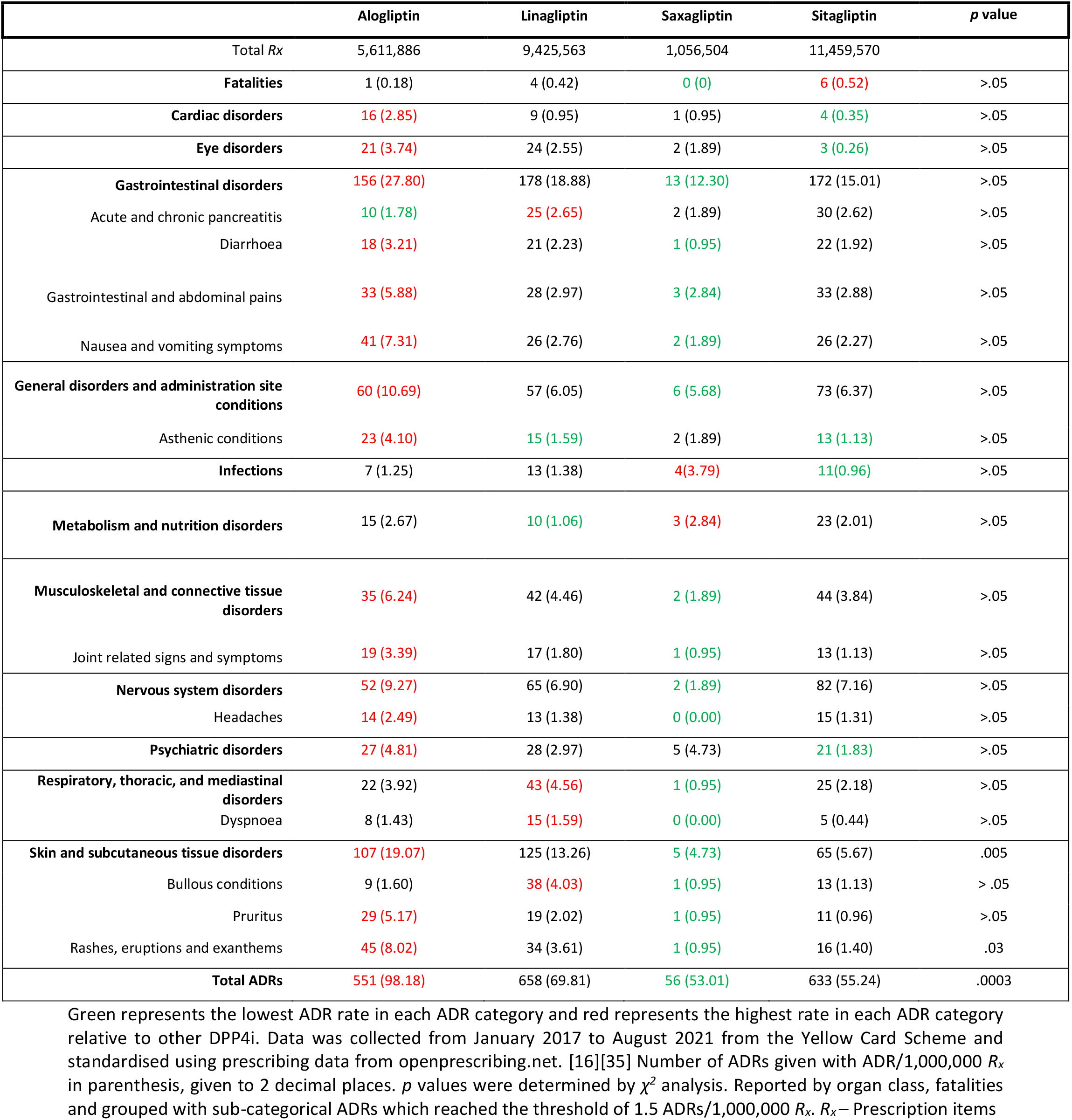
Adverse Drug Reactions for the DPP4i studied

### ADR Summary

Alogliptin had the highest ADR rate in cardiac, eye, gastrointestinal, general, musculoskeletal, and connective tissue, nervous system, psychiatric and skin disorders. In cardiac ADRs, alogliptin (2.85) showed a 3-fold increase on the next highest rate (0.95 for linagliptin and saxagliptin, respectively) and an 8-fold difference over sitagliptin (0.35); however, this did not reach a statistical difference. Alogliptin reached statistical difference against sitagliptin in skin disorders and saxagliptin in gastrointestinal, nervous, and skin disorders. Saxagliptin consistently showed the lowest ADR/1,000,000 *R*_*x*_ rate for most ADR types except for metabolism and nutrition disorders (2.84) and infections (3.79), where it showed the highest rate across the four drugs. Sitagliptin showed the highest fatality rate (0.52) and saxagliptin the lowest with no recorded fatalities; however, this did not reach statistical difference.

Overall, the highest rated ADRs across the class were gastrointestinal and skin disorders. In both organ classes, alogliptin experienced the highest rates (27.80 and 19.07, respectively), followed by linagliptin (18.88 and 13.26), sitagliptin (15.01 and 5.67) and saxagliptin (12.30 and 4.73).

## Discussion

The dipeptidyl-peptidase (DPP) family of enzymes includes homologous enzymes DPP2, DPP4, DPP8, DPP9 and FAP and are all *N*-terminal dipeptide cleaving serine proteases with preferential action where the second amino acid is proline or alanine where the most commonly seen multitarget interactions of the DPP4i’s studied (**Table 2**). [4][5][36] These proteins are known as DPP4 activity and/or structure homologue (DASH) proteins.[37] All members of the family have physiological processes associated with the immune system and inflammation, [38] which is attributed to their similar mechanisms of action and similar structures. Whilst DPP4 and FAP are extracellular, DPP8 and DPP9 are intracellular, specifically localised to the nucleus and cytosol, [38][39] which may make their function more redundant than that of DPP4. The exact physiological functions of DPP8 and DPP9 have yet to be fully elucidated. [37] DPP4 is both a type II transmembrane protein and has a soluble form which lacks the cytoplasmic tail and transmembrane region which circulates in plasma. [36] DPP4 is ubiquitously expressed, with particularly high expression in the kidney, lung, liver, and small intestine, [37] as well as circulating enzymatic activity as the cleaved domain continues to exert the effects in serum.

Notably, the *C*_max_ for all four drugs are too low to reach the necessary concentrations to inhibit off-target proteins such as DPP8, DPP9 and Fibroblast Activation Protein (FAP). This might explain why the suspected ADR rates across the drug class are not large or statistically different as the DPP4i class have similar pharmacological actions. The only clinically likely IC_50_ relative to the C_max_ would be linagliptin interacting with FAP and M1 and saxagliptin with DPP8 and DPP9, although there is a large discrepancy between the two values. Potential accumulation of drug might occur through decreased excretion or metabolism or accumulation over multiple doses depending on the half-life of the drug. [40]

Clinical trials identified gastrointestinal, skin and infections as common/uncommon ADRs of DPP4i (**S4**). DPP4i have a well-established association with these two ADRs (**S1**) [41] further confirmed by our study of the Yellow Card Scheme. Our results have confirmed that these are the most reported ADRs and found minimal statistical difference between the drugs in the class.

### Pharmacokinetic and physiochemical properties in relation to pharmacological data and ADRs

All five DPP4i were designed around the two basic amino acids; proline and alanine, giving the four DPP4i some structural similarity,[42] (**Figure 1**). This in turn generates similar PK and physiochemical properties (**Table 1**). Notably, linagliptin was the most lipophilic and had the strongest binding interaction with DPP4 and FAP (FAP exhibits the highest sequence identity to DPP4 of the DASH proteins).[37] Regardless of binding affinity with DPP4, there appears to be no correlation with the reported ADR data.

### Gastrointestinal (GI)

One notable ADR across the DPP4i class is acute and chronic pancreatitis with the highest rate associated with linagliptin (2.65 ADRs/1,000,000 *R*_*x*_) and lowest rate with alogliptin (1.78). Acute pancreatitis in DPP4i therapy warranted an MHRA alert (2014) due to the risk identified via pharmacovigilance.[43]

One possible underlying mechanism is the increased incretin hormone GLP-1 circulation-time which induces overgrowth of pancreatic acinar and ductal cells,[44][45] causing occlusion and resulting in pancreatitis;[46] although it must be noted that *β*-cell proliferation was induced with physiologically unlikely concentrations of GLP-1 and there is conflicting evidence to a significant association of pancreatitis with DPP4i therapy.[47][48][49][50]

DPP8 and DPP9 are more closely associated with gut inflammation and colitis.[51][52] Saxagliptin has the highest interaction with DPP8 and DPP9 (242 nM and 102 nM respectively) and exhibits the fewest total GI ADRs and sub-categorical GI ADRs. This may be attributed to the decreased inflammation associated with DPP8 and DPP9 resulting in less local irritation causing ADRs such as diarrhoea, abdominal pains and nausea and vomiting, for which saxagliptin had the fewest reported side effects across the class. Another suggested mechanism is due to the prolonged anti-motility activity of GLP-1.[41]

### Skin

DPP4 expression is ubiquitous in the dermis,[53] and the inhibition of DPP4 contributes to several cutaneous conditions, such as psoriasis,[52][54] atopic dermatitis,[55] cutaneous T-cell lymphoma and keloids,[53] amongst others.[56]

One rare but notable adverse reaction of DPP4i, is the development of bullous pemphigoid (BP),[41][57][58] an autoimmune cutaneous disease with uncertain pathogenesis. Histological features of BP vary depending on whether it is DPP4i-induced or of classical pathogenesis.[53] DPP4 has an established role in converting plasminogen to plasmin; one suggested mechanism is that DPP4i prevents plasmin cleavage of collagen XVII,[53][59] resulting in the breakdown of immunotolerance against collagen and production of autoantibodies against distinct collagen epitopes. Furthermore, enhancement of the activity of proinflammatory chemokines, such as CCL11/exotoxin, results in blister formation,[59] as well as a possible interference with keratinocyte migration and delayed wound healing.[60]

Linagliptin has the highest rate of bullous conditions (4.03) which may be attributed to its moderate affinity with FAP (89 nM); whereas other DPP4i affinity with FAP is significantly weaker. FAP has a defined role in collagen cleavage,[37] which also promotes macrophage adhesion,[61] and therefore the inhibition of FAP may lead to an additional autoantibody-mediated response as well as less macrophage adhesion and activity, which impacts the formation of blisters and skin healing.

### Immunological role (joints and infection)

DPP4, also known as CD26, is involved in amplifying the co-stimulatory signalling required for T-cell receptor activation and subsequently has an immune modulatory function.[37] Increased DPP4 activity is associated with decreased severity of rheumatic disease due to a dual role of CD26 involving cytokine inhibition and inducing cellular immunity.[36] It is logical therefore that DPP4i are associated with increased arthralgia and joint stiffness,[37] to the extent that an U.S. Food and Drug Administration (FDA) alert (2015) was circulated highlighting the association between DPP4i and severe and disabling joint pain.[62] Alogliptin was reported to have the highest ADR rates associated with musculoskeletal disorders (6.24) and joint signs and symptoms (3.39) and saxagliptin reported to have the lowest (1.89 and 0.95, respectively). This could be attributed to the increased affinity of saxagliptin with DPP8/9 which augment the immune modulatory processes of DPP4 and consequently exacerbate rheumatic symptoms.

By supressing T-lymphocyte-activating cytokines, chemokines and peptide hormones, there is an increased risk of infections[41][63]; namely nasopharyngitis, sinusitis, upper respiratory tract infections and urinary tract infections.[5][64] Furthermore, the potential inhibitory action of DPP8 and DPP9, which also has known T-lymphocyte-activating effects, may increase the immune modulation and increase the risk of infections.

Saxagliptin had the highest rate of infection (3.79) compared to linagliptin (1.38), alogliptin (1.25) and sitagliptin (0.96) respectively. This may be attributed to the increased affinity for the DPP8, DPP9 and FAP enzymes which, as discussed, all replicate and exaggerate the immunological function of DPP4.[65] This accumulation of anti-T-lymphocyte activation may cause the increased risk of infections; however, we would expect a correlating increase in joint pain or symptoms for saxagliptin if this were the case, although saxagliptin experienced the lowest musculoskeletal ADRs.

### Cardiovascular

Whilst alogliptin appears to exhibit a difference in cardiac ADR rate over sitagliptin, statistical significance across the DPP4i class was not reached. An FDA safety review has found that type 2 diabetes medicines containing saxagliptin and alogliptin may increase the risk of heart failure, particularly in patients who already have heart or kidney disease.[66] These findings were replicated in previous studies investigating the cardiac safety of alogliptin in type II diabetes, [50][67] which concluded that alogliptin had no additional cardiovascular risk than the placebo. Further trials have conferred that each DPP4i has no additional cardiovascular risk or excessive cardiac toxicity profiles, [47][48][49][50][68] which is particularly important given that patients with type II diabetes are at two-to four-fold increased risk of cardiovascular events. [50] Furthermore, it has also been postulated that this class exerts cardiovascular protection through GLP-1 modulation *in vivo*, which has well established cardiovascular effects through coronary artery endothelial cell proliferation and vasculoprotective endothelial progenitor cell stimulation.[69][70] Acknowledging that DPP4i increase GLP-1 levels, known benefits of GLP-1 should theoretically be augmented with DPP4i therapy. Furthermore, DPP4i can reduce pro-inflammatory cytokines, such as MCP-1,[71] which is associated with atherosclerotic plaques and visceral fat, which both contribute to immune-inflammatory disease, closely associated with diabetes. Reduction in chronic, low-grade inflammation could result in reduced cardiovascular pathology in type II diabetes. This could explain why the least reported organ class was cardiac disorders and shows DPP4i are associated with minimal cardiovascular toxicity. Whilst there is a theoretical benefit, the major cardiovascular trials for each DPP4i did not confer any cardiovascular benefit in a large-scale study. [47][48][49][50][68]

### Total ADRs

From visual inspection of the ADR rates and IC_50_ inhibition, there is little correlation between selectivity for DPP4, off-target interactions and total ADRs. Whilst linagliptin had the strongest interaction (1 nM), the total ADR rate for linagliptin was the second highest of all four drugs, suggesting that there is no relationship between strong on-target interactions and reduced ADRs. Saxagliptin had the lowest ADR rate but only the second lowest IC_50_, further indicating little correlation between these factors. This may be attributed to the similar physiological functions across the DPP4 family meaning that on-target ADRs and other interactions, with varying strengths of interaction, play a role in the overall toxicity profile of a drug.

## Limitations

Vildagliptin did not fit the scope for data availablity and was omitted from analysis. The Yellow Card Scheme relies on patients and healthcare professionals (HCPs) to report a ‘suspected’ adverse reaction of a drug therapy; however, causality does not need to be demonstrated and there is limited information about pre-existing co-morbidities, polypharmacy, genetics, or other factors.[10] Some ADRs may be a result of confounding factors, such as comorbidities, which cannot be identified from the Drug Analysis Profiles. Furthermore, time pressures on HCPs can lead to under-reporting of ADRs.[72]

Further factors can influence reporting of ADRs such as media reports or MHRA alerts which may result in increased reports for a period thereafter. The Weber effect is a well-observed effect where reporting reduces dramatically one-year post-approval (black triangle status of new drugs);[73] considering DPP4i have been licensed for at least 8 years, it is likely that ADRs are now under-reported. Moreover, patients may stratify side effects in terms of severity and consider some types more concerning than others, leading to higher reporting rates than other ADRs considered less important to report. [17][74]

Post-marketing surveillance systems also do not quantify the severity of an ADR; for example, one drug might cause a more severe headache than another, but this is not evaluated by the reporting system and therefore cannot be discussed in this study.

The Yellow Card Scheme is a UK-wide spontaneous reporting system; however, as openprescribing.net is English primary care only, the nationwide ADR rates may appear higher than the true value. OpenPrescribing.net measures the number of items prescribed; however, offers limited information regarding tablet strength or dose prescribed and therefore, the number of items prescribed may not necessarily equal to the number of patients. **S3** shows the tablet strengths available for each DPP4i and therefore if multiple tablets are prescribed to make the BNF indicated dose, ADRs may appear higher than the true value (the exception being linagliptin which has only one formulated strength tablet).

Whilst standardisation is often ADRs/100,000 *R*_*x*_,[19][75] the resulting numbers were not suitable for Chi squared analysis (which is only valid where numbers are >5).[76] Therefore, standardised the ADRs per million prescriptions improve the validity; however, it should be noted that any significant difference in the data reported relates to a population level risk rather than an individual risk.

Although saxagliptin is not licensed as a patch in the UK, the Yellow Card Scheme profile for saxagliptin listed transdermal patch as a formulation. These side effects were included in this study; however, it may lead to a higher rate of skin reactions in comparison to other DPP4i which could not be mitigated against.

## Conclusions

We have demonstrated that DPP4i have low suspected ADR rates and little statistical difference in types of ADR or differences in pharmacology. Whilst alogliptin appeared to demonstrate more ADRs in most organ classes, which may require further monitoring, this only reached statistical significance in total ADRs and other sub-categories such as nervous system (against saxagliptin) and skin (against saxagliptin and sitagliptin).

Saxagliptin, the least prescribed drug, consistently showed the lowest ADR rates in reported classes; again, rarely reaching significance showing the uniformity in ADR profile across the four DPP4i. This study was able to confirm that the most common experienced ADRs with this class were the already-established GI and skin reactions.[41]

We have postulated underlying mechanisms for these ADRs in relation to their pharmacological, physiochemical, and pharmacokinetic profiles.

The results demonstrate that the four DPP4i investigated have very similar structural and pharmacokinetic profiles, except for linagliptin, which showed increased lipophilicity and altered pharmacokinetic activity. Regardless of structural differences, the pharmacological activity against DPP4 did not correlate with the total suspected ADRs.

The non-DPP4 targets of the DPP4i’s of possible physiological relevance were identified as DASH proteins and therefore homologues of the desired protein target, meaning most off-target effects were an augmentation of DPP4-mediated adverse effects. Namely DPP8, DPP9 and FAP, with their role in immune modulation, or the prolonged action of incretin hormones GIP and GLP-1.

Further monitoring of pharmacovigilance schemes and a 5-year review of this data using this methodology may identify statistical difference between the DPP4is. Continuing monitoring and further research into the cardiac toxicity profile may determine whether there is a protective effect or cardiac damage caused from this therapy.

## Data Availability

All underlying data can be found in the supporting materials.

## Acknowledgements

We thank ChEMBL, MHRA Yellow Card Scheme and Open Prescribing for providing publicly available data.

## Data Availability Statement

All underlying data can be found in the supporting materials.

