## Supplementary Information for "Suspected Adverse Drug Reactions of the Type 2 Antidiabetic Drug Class Dipeptidyl-Peptidase IV inhibitors (DPP4i): Can polypharmacology help explain?"

**CONTENTS**

**S1.** Drug vs Drug Chi Squared ($\chi^2$) analysis. PAGE 2

**S2.** Full list of median $IC_{50}$ values (nM) for four DPP4i against named proteins. PAGE 3

**S3.** Indicated dose and formulated table strength of DPP4i PAGE 4

**S4** ADR findings of Phase III controlled clinical trials. PAGE 5

**S1.** Drug vs Drug Chi Squared ($\chi^2$) analysis.

| | ALOvsLINA | ALOvsSAXA | ALOvsSITA | LINAvsSAXA | LINAvsSITA | SAXAvsSITA |
|---|---|---|---|---|---|---|
| **Fatalities** | 0.75 | 0.67 | 0.68 | 0.51 | 0.92 | 0.47 |
| **Cardiac disorders** | 0.33 | 0.33 | 0.16 | 1.00 | 0.60 | 0.60 |
| **Eye disorders** | 0.63 | 0.44 | 0.082 | 0.76 | 0.17 | 0.27 |
| **Gastrointestinal disorders** | 0.19 | 0.014 | 0.05 | 0.24 | 0.51 | 0.60 |
| Acute and chronic pancreatitis | 0.68 | 0.95 | 0.69 | 0.72 | 0.99 | 0.73 |
| Diarrhoea | 0.67 | 0.27 | 0.57 | 0.47 | 0.88 | 0.57 |
| Gastrointestinal and abdominal pains (excl oral and throat) | 0.33 | 0.30 | 0.31 | 0.96 | 0.97 | 0.99 |
| Nausea and vomiting symptoms | 0.15 | 0.074 | 0.10 | 0.69 | 0.83 | 0.85 |
| **General disorders and administration site conditions** | 0.26 | 0.22 | 0.30 | 0.91 | 0.93 | 0.84 |
| Asthenic conditions | 0.29 | 0.37 | 0.20 | 0.87 | 0.78 | 0.66 |
| **Infections** | 0.94 | 0.26 | 0.85 | 0.29 | 0.78 | 0.19 |
| **Metabolism and nutrition disorders** | 0.40 | 0.94 | 0.76 | 0.37 | 0.59 | 0.71 |
| **Musculoskeletal and connective tissue disorders** | 0.59 | 0.13 | 0.45 | 0.31 | 0.83 | 0.42 |
| Joint related signs and symptoms | 0.49 | 0.24 | 0.29 | 0.61 | 0.70 | 0.90 |
| **Nervous system disorders** | 0.56 | 0.027 | 0.60 | 0.09 | 0.94 | 0.080 |
| Headaches | 0.57 | 0.11 | 0.54 | 0.24 | 0.97 | 0.25 |
| **Psychiatric disorders** | 0.51 | 0.98 | 0.25 | 0.53 | 0.60 | 0.26 |
| **Respiratory, thoracic and mediastinal disorders** | 0.83 | 0.18 | 0.48 | 0.12 | 0.36 | 0.48 |
| Dyspnoea | 0.92 | 0.23 | 0.47 | 0.21 | 0.42 | 0.51 |
| **Skin and subcutaneous tissue disorders** | 0.31 | 0.0033 | 0.0071 | 0.044 | 0.081 | 0.77 |
| Bullous conditions | 0.31 | 0.68 | 0.78 | 0.17 | 0.20 | 0.90 |
| Pruritus | 0.24 | 0.088 | 0.089 | 0.53 | 0.54 | 0.99 |
| Rashes, eruptions and exanthems | 0.20 | 0.018 | 0.031 | 0.21 | 0.32 | 0.77 |
| **Total ADRs** | 0.029 | 0.00024 | 0.00053 | 0.13 | 0.19 | 0.83 |

*Those with <0.05 are considered statistically different.*
*Values given to 2 significant figures.*

**S2.** Full list of median $IC_{50}$ values (nM) for four DPP4i against named proteins.

| | Alogliptin benzoate | Linagliptin | Saxagliptin hydrochloride | Sitagliptin phosphate |
|---|---|---|---|---|
| DPP4 | 5.3 | 1 | 3.385 | 18 |
| DPP8 | 100000 | 70000 | 242 | 48000 |
| DPP9 | 100000 | 100000 | 102 | 100000 |
| FAP | 100000 | 89 | 1000 | 100000 |
| M1 | | 297.5 | | |
| CYP2C19 | 10000 | | | |
| CYP1A2 | 10000 | | | |
| CYP2C9 | 10000 | | | |
| CYP2D6 | 10000 | | | |
| CYP3A4 | 20000 | | 100000 | |
| DPP7 | 100000 | 100000 | 30000 | 100000 |
| PREP | 100000 | 100000 | | 100000 |
| ERG | | 30000 | | |
| ACE | | | | 11000 |
| APN | | | | 100000 |
| NEP | | | | 100000 |
| PPCE | | | | 100000 |

| | |
|---|---|
| | 1-10 nM |
| | 10-100 nM |
| | 100-1000 nM |
| | 1000-10000 nM |
| | Weak inhibition (>10000 nM) |
| | Undetermined |

The Cmax (nM) of each DPP4i is provided to compare physiological relevance. Only homosapien, single protein assays were included in the averaged data. Key shows colour which correlates to stated strength interaction. Proteins: DPP – Dipeptidyl peptidase; FAP – Fibroblast Activation Protein; M1 – Muscarinic acetylcholine receptor 1; CYP – Cytochrome P450.

**S3.** Indicated dose and formulated table strength of DPP4i

| | Alogliptin | Linagliptin | Saxagliptin | Sitagliptin |
|---|---|---|---|---|
| BNF Indicated dose | 25mg OD | 5mg OD | 5mg OD | 100mg OD |
| Formulated tablet strengths | 6.25mg<br>12.5mg<br>25mg | 5mg | 2.5mg<br>5mg | 25mg<br>50mg<br>100mg |

**S4** ADR findings of Phase III controlled clinical trials.

| | Alogliptin | Linagliptin | Saxagliptin | Sitagliptin |
|---|---|---|---|---|
| **Infections**: | | | | |
| Upper respiratory tract | Common | - | Common | |
| Urinary tract infection | - | | Common | |
| Nasopharyngitis | Common | Uncommon | - | |
| Gastroenteritis | - | - | Common | |
| Sinusitis | - | - | Common | |
| **Gastrointestinal**: | | | | |
| Pancreatitis | Not Known [a] | Rare [b] | Uncommon [a] | Not known [a] |
| Abdominal pain | Common | - | Common [a] | |
| Diarrhoea | - | | Common | |
| Constipation | | | Not known [a] | Uncommon |
| Nausea | | | Common [a] | |
| Vomiting | | | Common | Not known [a] |
| Gastroesophageal reflux disease | Common | | | |
| Hepatic dysfunction | Not Known [a] | | | |
| **Nervous system**: | | | | |
| Dizziness | - | - | Common | Uncommon |
| Headache | Common | | Common | Common |
| Fatigue | | | Common | |
| **Skin**: | | | | |
| Pruritus | Common | | Uncommon [a] | Uncommon [a] |
| Rash | Common | Uncommon [a] | Common [a] | Not known [a] |
| Erythema multiforme | Not Known [a] | | | |
| Angioedema | Not Known [a] | Rare [a] | Rare [a] | Not known [a] |
| Urticaria | Not Known [a] | Rare [a] | Uncommon [a] | Not known [a] |
| Bullous pemphigoid | - | Rare [b] | Not known [a] | Not known [a] |
| Dermatitis | | | Uncommon [a] | |
| **Other:** | | | | |
| Cough | - | Uncommon | | |
| Thrombocytopenia | | | | Rare |
| Amylase increased | | Uncommon | | |
| Lipase increased | | Common | | |
| Hypersensitivity | Not Known [a] | Uncommon | Uncommon [a] | Not known [a] |
| Anaphylaxis | | | Rare [a] | |

Frequencies are defined as very common (≥1/10); common (≥1/100 to <1/10); uncommon (≥1/1,000 to <1/100); rare (≥1/10,000 to <1/1,000); very rare (<1/10,000); not known (cannot be estimated from available data). Data collected from European Medicines Agency summary product characteristics.[a] Observed from post-marketing surveillance; [b] based on CARMELINA trial
